# Diffusion kurtosis imaging of white matter in bipolar disorder

**DOI:** 10.1101/2021.02.01.21250951

**Authors:** Vina M Goghari, Mavis Kusi, Mohammed K Shakeel, Clare Beasley, Szabolcs David, Alexander Leemans, Alberto De Luca, Louise Emsell

## Abstract

**Objectives:** White matter pathology is thought to contribute to the pathogenesis of bipolar disorder (BD). However, most studies of white matter in BD have used the simple diffusion tensor imaging (DTI) model, which has several limitations. DTI studies have reported heterogenous results, leading to a lack of consensus about the extent and location of white matter alterations. Here, we applied two advanced diffusion magnetic resonance imaging (MRI) techniques to investigate white matter microstructure in BD.

**Methods:** Twenty-five patients with BD and 24 controls comparable for age and sex were included in the study. Whole-brain voxel-based analysis (VBA) and a network-based connectivity approach using constrained spherical deconvolution (CSD)-tractography were used to assess group differences in diffusion kurtosis imaging (DKI) and DTI metrics.

**Results:** VBA showed lower mean kurtosis in the corona radiata and posterior association fibers in BD following threshold-free cluster enhancement. Regional differences in connectivity were indicated by lower mean kurtosis and kurtosis anisotropy in streamlines traversing the temporal and occipital lobes, and lower mean axial kurtosis in the right cerebellar, thalamo-subcortical pathways in BD. Significant differences were not seen in the DTI metrics following FDR-correction.

**Conclusions:** Differences between BD and controls were observed in DKI metrics in multiple brain regions, indicating altered connectivity across cortical, subcortical and cerebellar areas. DKI was more sensitive than DTI at detecting these differences, suggesting that DKI is useful for investigating white matter in BD.

## Introduction

Accumulating evidence from neuroimaging studies suggests that white matter pathology plays a role in the pathogenesis of bipolar disorder (BD). White matter abnormalities in BD were first observed as increased rates of white matter hyperintensities in anatomical magnetic resonance imaging (MRI) scans.^1^ Further evidence supporting the possibility of white matter abnormalities in BD has been provided by several voxel-based morphometry (VBM) studies that have reported reductions in white matter density and volume in people with BD compared to controls.^2^ Moreover, diffusion MRI (dMRI) studies have often reported observations of white matter abnormalities in BD.^3,4^

To date, the most commonly applied model in studies of white matter microstructure in BD is the diffusion tensor imaging (DTI) model. DTI metrics include fractional anisotropy (FA), mean diffusivity (MD), axial diffusivity (AD), and radial diffusivity (RD). The DTI metric, FA is a measure of the degree to which water diffusion is constrained in the brain.^5^ FA is often used as a general index of axonal integrity, although this interpretation is an over-simplification as FA is also sensitive to other biological and technical parameters, see Jones et al.^6^ Damage to white matter or demyelination along axons results in water movement being more isotropic, which may manifest in relatively low FA values. While DTI studies have generally reported lower FA in BD patients compared to controls,^7–9^ some studies have found FA values to be higher^10,11^ or similar between groups.^12^ These inconsistencies may reflect heterogeneity in methodology, or sample characteristics such as age of onset, psychotic features, disease duration, and medication type, or may result from small sample sizes. Similarly, there is a lack of consensus as to the location of white matter abnormalities in BD. One of the first whole-brain DTI studies^11^ identified white matter alterations predominantly in frontal regions in BD, while numerous DTI studies that followed found evidence of alterations in frontal and limbic areas ^3,4^, consistent with neural models of BD that suggest that fronto-limbic dysconnectivity underlies symptoms of this disorder.^13^ However, a number of whole-brain DTI studies have reported white matter abnormalities in BD to be widespread.^7,8,14^

An important limitation of most DTI studies in BD is that they only use FA and fail to utilize other valuable DTI metrics such as MD, AD and RD. Alterations in specific white matter characteristics cannot be identified by analyzing changes in FA alone;^3^ therefore, investigating additional DTI metrics may provide complementary information. MD reflects the average molecular motion, independent of tissue directionality and is usually higher in damaged tissue.^15^ AD and RD, respectively refer to the degree of diffusion parallel and perpendicular to the main direction of diffusion. RD is typically increased in areas that show reduced FA and may indicate disrupted myelination.^15^ Few studies have analyzed MD, AD and RD in BD patients, and results have been mixed, with some studies finding these DTI metrics to be lower,^16,17^ higher ^5,7^ or unchanged^5,18^ in BD. Further studies are needed to elucidate the association between BD and these DTI metrics.

Despite its widespread use, DTI has several limitations. Most notably, DTI assumes the presence of a single type of diffusion in each imaged voxel, which limits its specificity as any observed change could be ascribed to either intra or extra-axonal water. Moreover, DTI cannot distinguish between the orientation of different fiber bundles within a voxel, so called “crossing fibers”, and hence cannot reliably reconstruct fiber tracts through regions with more than one fiber population, i.e., the majority of the brain.^19^ More recently, novel models address some of the limitations of DTI, while increasing the specificity and sensitivity to properties of the in-vivo diffusion process. One such model is diffusion kurtosis imaging (DKI). DKI extends the DTI model by quantifying the amount of non-Gaussian diffusion and is thus more sensitive to restricted diffusion within cells.^20^ DKI provides several analogous, but complementary quantitative measures to DTI, which are based on the kurtosis tensor: kurtosis anisotropy (KA), mean kurtosis (MK), axial kurtosis (AK), and radial kurtosis (RK). Of note, while DKI and DTI metrics are based on a similar tensor framework, they are not equivalent, nor well correlated in neural tissue, thus reflecting the ability of DKI metrics to capture different information to DTI measures. For example, while MD is sensitive to average differences in extracellular diffusion, MK reflects differences in both intracellular and extracellular compartments and provides an indirect measure of tissue complexity; MK is generally higher for more complex or heterogenous microstructures.^21^

While DTI can be used to perform fiber tractography, it is not an optimal technique as it cannot reliably reconstruct fiber tracts through regions of crossing fibers. Constrained spherical deconvolution (CSD) tractography, based on high angular resolution dMRI data, has been proposed as a more reliable alternative for fiber tractography. CSD differs from the tensor model by estimating a fiber orientation density function (fODF) within each voxel, whose width and orientation provide an estimate of the local distribution and density of fiber populations.^22^ Unlike the diffusion tensor, which can only model a single orientation, the fODF can distinguish multiple fiber populations within each voxel with commonly employed acquisitions. This means that CSD based tractography is capable of tracking through regions of crossing fibers, which makes it particularly suited to studying structural connectivity.^22^

The main objectives of the current study were three-fold: First, to investigate the presence of differences in DKI metrics between people with BD and community controls using two complementary dMRI techniques: a whole-brain voxel-based analysis (VBA) and a network- based connectivity analysis based on CSD tractography. Second, to investigate whether DKI is more sensitive to white matter alterations in BD than DTI parameters. Third, to explore the relationship between any differences in dMRI measures and mood, cognition, and global functioning in the BD cohort. We hypothesized that there would be group differences in both DKI and DTI metrics in widespread brain regions, including lower MK and lower FA, reflecting altered tissue microstructure in these areas. Moreover, we anticipated that the DKI differences would be more pronounced and would be correlated with the degree of mood, cognition and functioning scores in BD patients.

## Methods

### Participants

Twenty-eight patients with bipolar disorder and 27 community controls were recruited for the study. The patients with BD were recruited through an outpatient mood disorder clinic, through the Organization for Bipolar Affective Disorder Society in Calgary, Alberta, and online and community advertisements. Controls were recruited through community advertisements. All participants were screened using the following exclusion criteria: 1) age less than 18 or greater than 60; 2) diagnosis of a substance related disorder in the last three months (excluding nicotine, caffeine and cannabis); 3) use of inhalants three or more times; 4) history of head injury with loss of consciousness for more than 30 minutes; and 5) a history of electroconvulsive therapy, epilepsy, seizures, stroke, diabetes, legal blindness, MRI contraindications, or any medical or neurological condition that would make it impossible to complete the study. Controls were also excluded from the study if they had a history of major depressive disorder, had used antipsychotic or antidepressant medication, or had personal or family history of a psychotic or BD. Written informed consent was obtained from all participants and the study was approved by the University of Calgary’s Conjoint Health Research Ethics Board (CHREB).

### Clinical and cognitive assessments

Presence of mood, psychotic, substance use, or anxiety disorder was determined using the Structural Clinical Interview for the Diagnostic and Statistical Manual of Mental Disorders – 5^th^ Edition (SCID-5).^23^ The SCID-5 was also used to determine if the patients had bipolar I or II disorder. The Young Mania Rating Scale (YMRS)^24^ and the Hamilton Depression Rating Scale (HAM-D)^25^ were used to evaluate recent symptoms of mania and depression, respectively. Functional ability was assessed using the Functioning Assessment Short Test (FAST)^26^ and the Social and Occupational Functioning Assessment Scale (SOFAS).^27^ Intelligence was estimated using the Wechsler Test of Adult Reading (WTAR).^28^

### Multi-shell diffusion MRI data acquisition

Images were acquired on a 3-tesla General Electric 750 MR scanner with a 12-channel receiving head coil. MRI data included a multishell diffusion-weighted image series, reversed-phase encoded diffusion-weighted acquisition, and a 3D T1-weighted image series. The multishell diffusion-weighted images consisted of 98 directions of diffusion weighting in total, distributed between 3 b-shells (b-values) as follows: 8 directions for b = 300 s/mm^2^, 30 directions for b = 700 s/mm^2^, and 60 directions for b = 2000 s/mm^2^ as well as 10 non-diffusion-weighted images (b = 0 mm^2^/s). The non-weighted images were distributed between the 3 different diffusion-weighted b shells (1, 3 and 6 per shell respectively). Images were acquired in the axial plane with the following parameters: isotropic voxel size = 2.5 mm, TR = 7250 ms, TE = 70 ms, in-plane parallel image acceleration with Array coil Spatial Sensitivity Encoding (ASSET factor = 2). The reversed-phase encoded diffusion series for echo-planar imaging (EPI) distortion correction, consisted of a set of 3 posterior-anterior (PA) diffusion-weighted images, i.e., one per diffusion weighted b-shell.

### Image pre-processing

Subject-motion, eddy current and susceptibility distortions were corrected using the FSL v.5.0.11 (https://fsl.fmrib.ox.ac.uk/fsl/fslwiki/FSL, RRID:SCR_002823)^29^ tools topup^30^ and eddy.^31^ First, the non-zero off-resonance fields were estimated from the b = 0 s/mm^2^ images with opposite phase encoding using 3 PA and 10 AP encoded volumes. The estimated fields were passed on to eddy. The following non-default settings were: eddy was forced not to check if data is shelled with the command “--data_is_shelled”, detection, and replacement of data with Gaussian Process predictions^31^ was performed with the command “-repol”. During motion correction, eddy rotated the DWI gradients^32^ as well. The brain mask for eddy was calculated from the mean unwarped images from the topup tool using BET (Brain Extraction Tool).^33^

### Image analysis and quantitative metrics

The dMRI data of each subject was processed with the ExploreDTI toolbox (http://www.exploredti.com, RRID:SCR_001643)^34^ for MATLAB R2018b (https://www.mathworks.com, RRID:SCR_001622) to derive quantitative metrics and study brain connectivity. The DKI model^35^ was fit to the dMRI data of each subject with a weighted-least squares approach^36^ while constraining the MK estimates within physiologically plausible values (0≤MK≤3).^20^ Subsequently, voxel-wise maps of MD, FA, AD, RD, MK, AK, RK, and KA were derived.

### Voxel-based analysis

To identify local differences in white matter between patients and controls, we performed a voxel-wise analysis of the DTI/DKI metrics. The FA map of each subject was registered with Elastix (https://elastix.lumc.nl, RRID:SCR_009619) ^37^ to the 1×1×1mm^3^ FA template of the FMRIB, which is part of FSL.^29^ To achieve fine voxel-wise alignment, each subject was registered with a combination of rigid, affine and b-spline transformations. Subsequently, the derived transformations were applied to co-register each metric to the template space. Voxel-wise t-tests controlling for age and sex were performed for each DTI/DKI metric testing the hypothesis CONTROLS > PATIENTS and PATIENTS > CONTROLS, separately. Statistics were performed with FSL “randomise” using 5000 permutations^38^ and the TFCE^33^ correction for multiple comparisons.

### Connectivity analysis

The CSD approach was fit to the data at b = 0, 2000 s/mm^2^ with ExploreDTI using the recursive calibration of the response function^39^ and spherical harmonics of order 8. Whole brain fiber deterministic tractography was performed using step-size equal to half the voxel-size and angle threshold of 45 degrees. The Destrieux atlas was aligned to the individual subject space by registering its template to the individual diffusion space with Elastix, then the connectivity matrices were derived accordingly. For each subject, the mean values of the DTI/DKI metrics between two nodes were derived, then significant differences between controls and patients were tested with the network-based statistics toolbox^40^ using a false-discovery-rate approach^41^ to correct for multiple comparisons.

### Associations between DKI metrics and mood, cognition and functioning in bipolar disorder

Finally, we explored whether microstructural differences between participants with BD and controls were associated with mood or neuropsychological function. Voxel-wise general linear model-based analysis using FSL “randomise” was utilized to investigate the relationship between any kurtosis measures that showed statistically significant voxel-wise group differences and FAST (total), SOFAS (total), YMRS (total), HAMD (total), WTAR (raw) scores in the BD group. The TFCE technique was employed to correct for multiple comparisons.

## Results

### Participant characteristics

Three BD patients and three controls were excluded due to poor image quality, leaving a final sample size of 25 BD subjects (23 bipolar I and 2 bipolar II patients) and 24 community controls. Nine of the patients included in the study met current criteria for bipolar I (four met criteria for a manic episode and five met criteria for a depressive episode) and one met current criteria for bipolar II (hypomanic episode). Participant characteristics are shown in Table 1. The BD and control groups did not differ by age, sex, handedness, or estimated intelligence.

**Table 1.** Demographic Characteristics for Bipolar Patients and Community Controls.

|  | Bipolar (n = 25) | Control (n = 24) | Test Statistics |
| --- | --- | --- | --- |
| Age (years): mean (SD) | 38.56 (11.62) | 38 (10.18) | $t(47) = 0.18, p = 0.86$ |
| Sex (% Female) | 51.4 | 48.6 | $X^2(1) = 0.008, p = 0.93$ |
| Education (Years Completed): mean (SD) | 14.42 (2.69) | 15.88 (1.92) | $t(41.62) = -2.16, p = 0.04$ |
| Handedness (% Right-Handed) | 96 | 91.7 | $X^2(2) = 1.07, p = 0.59$ |
| WTAR Standard Score: mean (SD) | 111.65 (11.70) | 109.52 (9.53) | $t(44) = 0.68, p = 0.50$ |
| YMRS: mean (SD) | 3.63 (3.75) | 0.83 (1.17) | $t(27.41) = 3.48, p = 0.002$ |
| YMRS: range (0-60) | 0-16 | 0-4 | - |
| HAM-D: mean (SD) | 7.38 (6.81) | 0.71 (1.30) | $t(24.68) = 4.71, p < 0.001$ |
| HAM-D: range (0-52) | 0-23 | 0-6 | - |
| FAST: mean (SD) | 12.29 (9.26) | 1.48 (2.25) | $t(25.83) = 5.55, p < 0.001$ |
| FAST: range (0-72) | 0-37 | 0-8 | - |
| SOFAS: mean (SD) | 66.18 (11.39) | 85.0 (7.45) | $t(27.07) = -5.79, p < 0.001$ |
| SOFAS: range (0-100) | 0-40 | 0-35 | - |
| Atypical Anti-Psychotics (% on) | 60 | 0 | - |
| Typical Anti-Psychotics (% on) | 8 | 0 | - |
| Anti-Convulsants (% on) | 56 | 0 | - |
| Anti-Depressants (% on) | 36 | 0 | - |
| Lithium (% on) | 28 | 0 | - |
| Anxiolytics (% on) | 4 | 0 | - |
| Sedative-Hypnotics (% on) | 8 | 0 | - |
| Other Psychiatric Medications (% on) | 4 | 4.2† | - |

WTAR, Wechsler Test of Adult Reading; YMRS, Young Mania Rating Scale; HAM-D, Hamilton
Depression Rating Scale; FAST, Functioning Assessment Short Test; SOFAS, Social and Occupational
Functioning Assessment Scale. †One participant in the control group was on lisdexamfetamine
which is mainly used to treat attention deficit hyperactivity disorder (ADHD); this participant's
SCID-5 scores did not show presence of a psychological disorder.

However, the two groups differed on years of education with controls having completed more years of education than the BD patients.

### Voxel-based analysis

VBA revealed that the BD patients had significantly lower MK than controls, mainly in the corona radiata and posterior association bundles. There were no regions where MK was lower in controls, nor were there any significant differences in any other DKI or DTI measures following TFCE correction for multiple comparisons (Figure 1). Maps thresholded at a less conservative threshold for the typically analysed metrics FA, MD and MK are provided as supplementary material for comparison.

**Figure 1.**
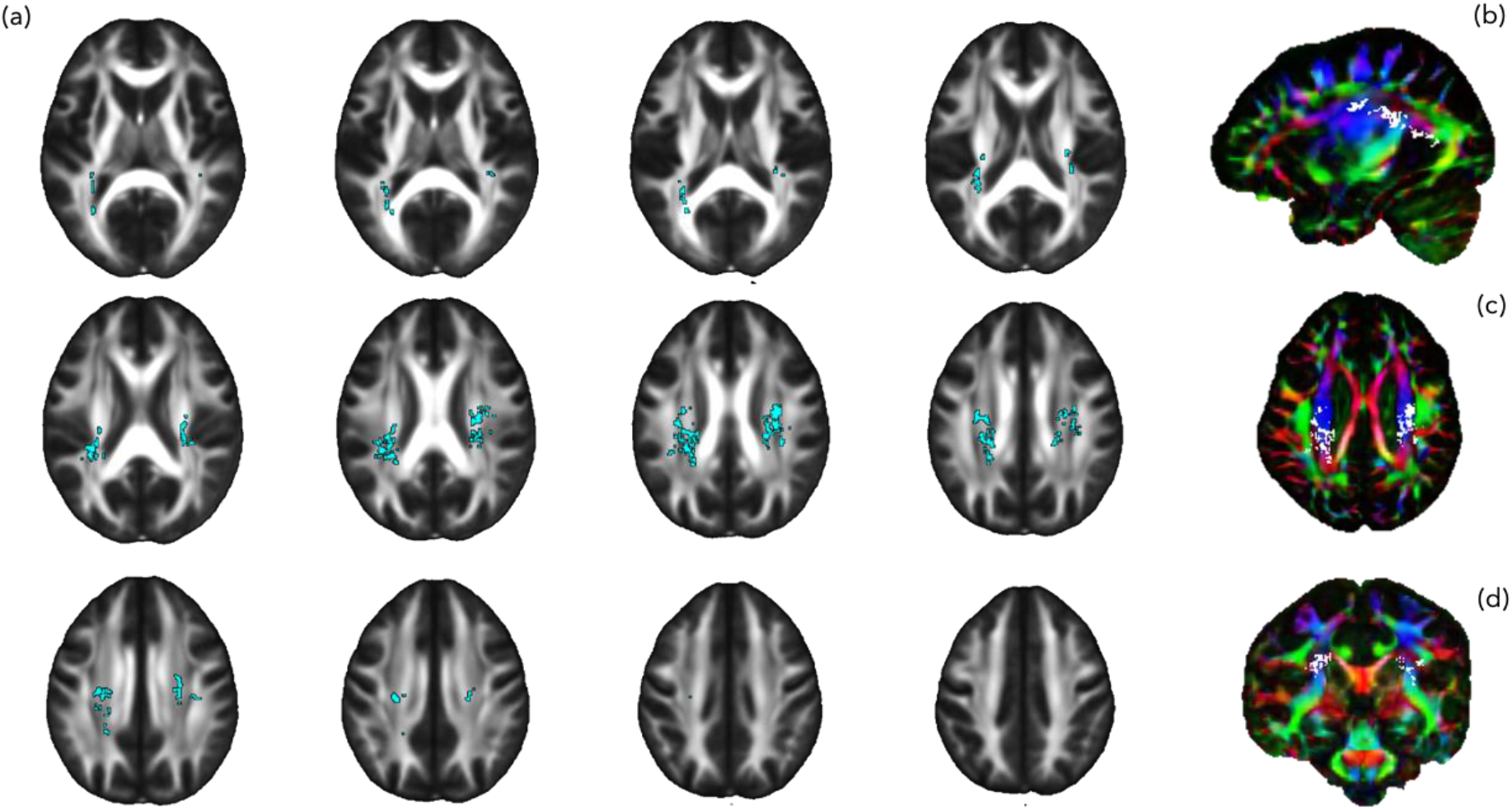
Voxel-wise differences in mean kurtosis (MK) between bipolar patients and controls Legend: (a) Voxel-wise binary map of regions where MK was significantly lower in individuals with bipolar disorder compared to controls following TFCE, FWE-corr p<.05, displayed on FMRIB58 FA template (a) multi-slice axial view. Results from (a) displayed on colour FA population in (b) sagittal, (c) axial and (d) coronal view for anatomical context. TFCE = threshold-free cluster enhancement, FWE = family-wise error

### Connectivity analysis

Regional differences in connectivity based on lower mean MK and KA were detected in connections traversing the temporal and occipital lobes, and lower mean AK was detected in right cerebellar, thalamo-subcortical pathways in the BD group. Specifically, significantly lower mean MK was detected in streamlines traversing the left lateral aspect of the superior temporal gyrus and left transverse temporal sulcus, and traversing the right angular gyrus and right supramarginal gyrus. Lower mean KA was detected in streamlines traversing the left superior occipital gyrus and left parieto-occipital sulcus. Lower mean AK was detected in streamlines traversing the brainstem and right thalamus, the right cerebellum and right thalamus, right pallidum and right precentral gyrus, and right cerebellar cortex and right central sulcus. There were no statistically significant group differences in DTI metrics. These results are displayed in Figure 2.

**Figure 2.**
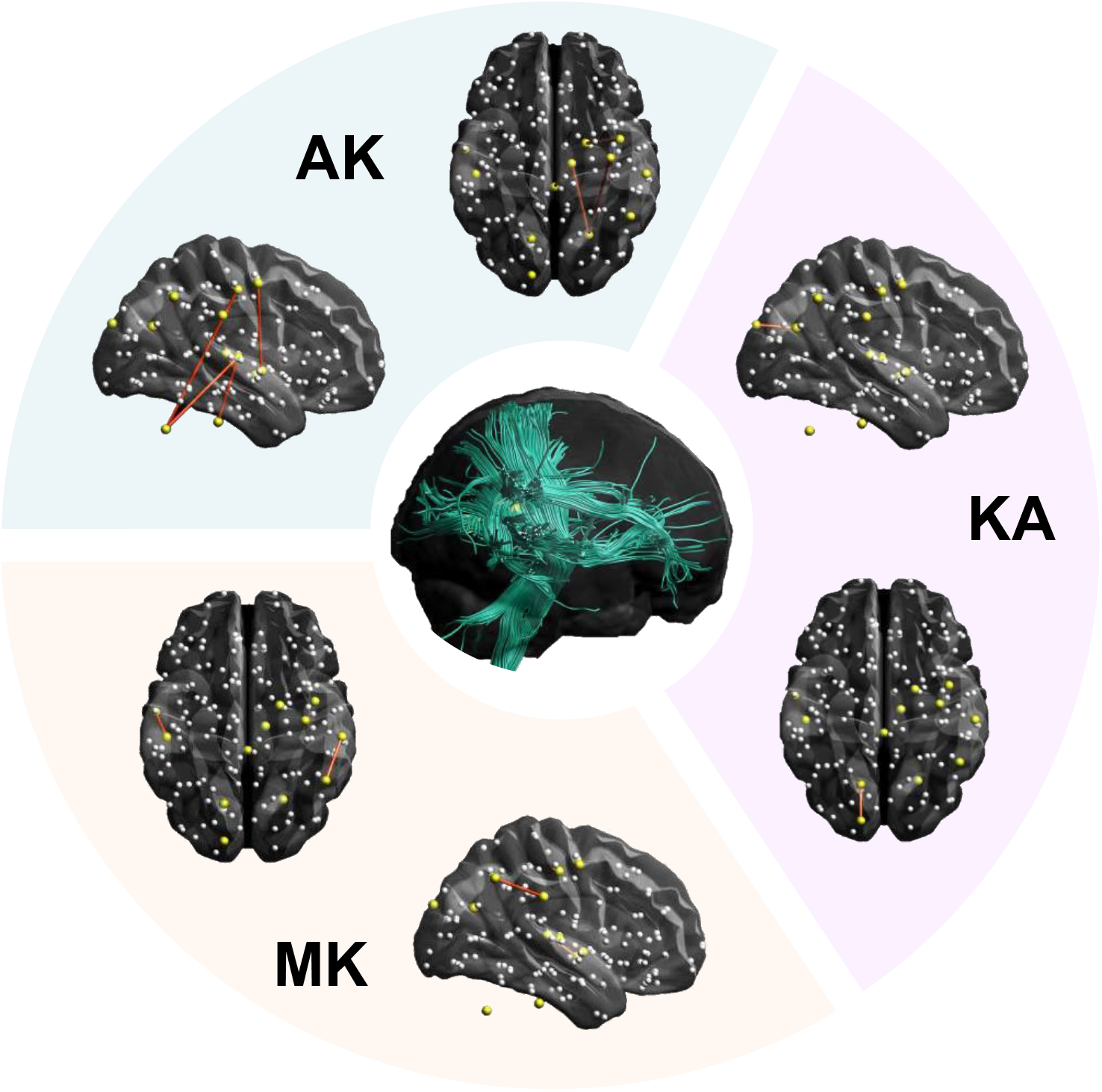
Tract-based differences in structural connectivity between bipolar patients and controls. Legend: Outer-circle: Significant inter-nodal group differences in mean DKI metrics between bipolar patients and controls represented by dark-red lines in axial and sagittal views. The yellow nodes were significantly different in at least one metric. The white nodes are the total tested nodes. Central figure: Connections involving streamlines (turquoise) traversing voxels with significantly different DKI metrics between bipolar patients and controls (yellow). DKI = diffusion kurtosis imaging.

### Associations between DKI measures mood, functioning and cognition in BD

There were no significant associations between any of the test scores (i.e., FAST, SOFAS, YMRS, HAMD, and WTAR) and MK, the only parameter that demonstrated a difference between groups.

## Discussion

The current study investigated microstructural changes in BD with DKI, using a whole-brain VBA and a network-based connectivity approach based on CSD tractography. To investigate possible BD related changes in white matter, we compared the DTI and DKI metrics of BD patients to those of controls. This analysis was also done to determine if DKI is more sensitive to white matter alterations in BD than DTI. We also examined possible associations between white matter changes in BD and mood and neuropsychological function. The VBA showed lower MK in BD patients compared to controls, predominantly in the corona radiata and posterior association fibers. There were no significant differences between the BD and control groups in any other DKI or DTI measures after applying conservative statistical thresholds, corrected for multiple comparisons. The CSD analysis revealed regional differences in connectivity as shown by lower mean MK and KA in connections traversing the temporal and occipital lobes, and lower mean AK in right cerebellar, thalamo-subcortical pathways in BD compared to controls. No significant associations between MK, the only parameter that differed between patients and controls in the VBA, and mood and neuropsychological function in the BD patients were observed in the current study.

In general, previous DTI studies of white matter in BD have reported significantly lower FA in BD patients relative to controls.^5,7–9,14,16^ Studying the largest sample size (*n* = 3033) to date, Favre et al.^8^ found significantly lower FA in widespread brain regions in BD patients, with the largest effect sizes in the whole corpus callosum, followed by the body and genu of the corpus callosum, and the bilateral cinguli. While not entirely consistent, DTI studies in BD that have examined other metrics have reported significant alterations in MD,^7^ AD^16,17^ and RD^5,16^ in this disorder. In contrast to these studies, we did not find significant changes in DTI measures in BD relative to the control group. This discrepancy could be due to differences in sample size, as studies reporting significant changes in DTI parameters in BD have tended to have larger samples.^7,8,14,16^ Of note, we applied conservative statistical thresholds to our analyses, and corrected for multiple comparisons; when applying a more liberal approach, we also detected differences in DTI measures (see supplementary material).

While significant differences were not detected in DTI metrics in our analyses, we detected significant differences in DKI parameters, in particular MK, between groups. These results suggest that DKI is more sensitive than DTI at detecting microstructural differences in BD. Our findings also suggest that applying DKI to study white matter in BD may prove to be more useful than DTI alone in pinpointing the location and extent of white matter alterations in BD. To date, few studies have used DKI to investigate white matter microstructure in BD. Ota et al.^42^ compared DTI and DKI metrics of BD patients and controls by means of voxel-wise analysis, finding significant reductions in FA in the bilateral cingulate cortices and significantly lower MK in the right inferior fronto-occipital fasiculus and the right posterior cingulate cortex in BD, relative to controls.^42^ Sawamura et al.^43^ examined microstructural alterations in BD compared to major depression and controls using whole-brain voxel-based DKI analysis. Significant microstructural alterations were found in widespread brain regions in BD, including lower KA in left temporal lobe, right superior frontal gyrus, right orbital gyrus, right cingulate gyrus, left insula, and right parahippocampal gyrus, compared to controls. The BD patients also had lower MK in the left frontal paraventricular white matter, right parietal lobe and right superior frontal gyrus and higher MK and RK in right precentral gyrus, compared to controls.^43^ The present study did not find such widespread differences in DKI parameters as the Sawamura et al.^43^ study, and revealed differences predominantly in posterior white matter tracts, whereas DKI differences in the Sawamura et al.^43^ study were located primarily within frontal brain areas. Methodological differences may contribute to the discrepancies between this study and that of the previous DKI studies in BD. Statistical analyses in the Ota et al.^42^ and Sawamura et al.^43^ studies were performed using SPM8 and SPM12, respectively, and they reported cluster-level uncorrected p-values < 0.001. While more stringent thresholds and multiple-comparison corrections were applied to the statistical analyses for this research. Differences in the DKI fit method might also account for the inconsistent findings; the DKI model was fit to the dMRI data using a weighted-least squares approach in this study, while Ota et al.^42^ and Sawamura et al.^43^ both used the Diffusional Kurtosis Estimator (DKE). Heterogeneity in the characteristics of the study participants, including BD subtype, age of onset, illness duration, and medication status might also account for inconsistencies between the studies. All three studies had relatively small sample sizes, indicating a need for DKI studies with larger samples.

Discrepancies were observed between VBA and CSD connectivity-based analysis in the present study. While MK was the only parameter to show significant group differences in the VBA, lower average MK and KA were identified in connections traversing the temporal and occipital lobes, and lower average AK was found in right cerebellar, thalamo-subcortical pathways in the connectivity analysis. Differences in results between VBA and connectivity-based analysis are likely due to differences in sensitivity. In this context, whilst replicating findings across methodologies is important for assessing their robustness, discrepancies between findings using different analysis approaches may be less problematic. For example, in our case, VBA and network analysis were used to test different hypotheses about the location and degree of microstructural difference in our study population, with the VBA assessing voxel-level differences as opposed to the more anatomically constrained whole-tract level of the connectivity analysis. It is therefore plausible that factors such as image registration and noise contribute differentially to the regional sensitivity of the two methods. The findings in this case are complementary and reveal that there is a high probability of non-specific microstructural alterations associated with BD. This forms a sound basis for future work applying multimodal imaging and other clinical data in larger samples to place the findings in context.

Few previous studies have used CSD tractography to investigate structural connectivity in BD. Emsell et al.^44^ conducted a ROI study to examine alterations in limbic and callosal white matter in euthymic BD type I patients. They found decreased FA and increased MD and RD in the left fornix, subgenual cingulum, and the whole corpus callosum in BD compared to controls, along with increased AD in the bilateral fornix and right dorsal-anterior cingulum.^44^ In a CSD tractography study that used whole-brain as well as ROI graph theory-based network analyses, Forde et al.^45^ found no differences between euthymic BD type I patients, first-degree unaffected relatives and controls in the whole-brain network analysis. However, the ROI analysis found evidence of impaired connectivity in the right medial superior frontal gyrus and left middle frontal gyrus in BD compared to controls.^45^

We did not find significant correlations between clinical or cognitive test scores and MK, which differed between patients and controls. This is consistent with several previous DTI and DKI studies that also failed to observe significant associations between clinical or cognitive test scores and diffusional indices.^9,18,42,44^ However, some DTI studies with large sample sizes have noted significant relationships between clinical variables and DTI parameters. In particular, Favre et al.^8^ reported a significant positive association between age at onset of BD and FA in the right inferior fronto-occipital fasiculus and a negative association between illness duration and FA in the left cingulum. Moreover, FA was significantly lower in the genu of the corpus callosum in patients receiving vs. those not receiving antipsychotics and in multiple ROIs in patients receiving vs. those not receiving anticonvulsants. Higher FA values were found in several regions in patients receiving vs. not receiving lithium.^8^ Furthermore, Koshiyama et al.^14^ observed significant negative correlations between duration of illness and FA in the fornix, anterior corona radiata, genu of the corpus callosum, and whole-brain white matter skeleton in BD patients. Additional studies with larger sample sizes might improve understanding of the correlation between DKI/DTI parameters and clinical variables in BD.

The observed changes in DKI metrics in the current study indicate the presence of white matter abnormalities in BD. In particular, reductions in MK and KA in the present study indicate loss of white matter microstructural complexity and altered connectivity in multiple brain regions. The reductions in AK reflect decreased microstructural complexity along the axial direction of white matter tracts^46^ in BD. White matter alterations in the corona radiata and posterior brain regions, including occipital white matter (e.g., posterior thalamic radiation, inferior longitudinal fasiculus, and inferior fronto-occipital fasiculus) have been reported in untreated first-episode BD with psychosis.^47^ This suggests that the changes that we observed in the corona radiata and posterior brain regions may appear early in the course of BD, are not related to psychotropic medications, and might be associated with psychotic symptoms. In line with our findings, white matter alterations have previously been reported in the cerebellum and other parts of the motor system.^16,48^ The disruptions in connectivity that we observed in motor regions might be associated with psychomotor slowing in BD. Bracht et al.^49^ investigated the white matter microstructure of motor regions in people with BD with a current episode of depression, finding a significant increase in FA in the left corticospinal tract of the BD patients. A positive association was observed between FA in the left corticospinal tract and activity levels in BD, indicating a link between white matter alterations in motor pathways and psychomotor slowing, which was interpreted as reflecting a compensatory role of the left corticospinal tract in psychomotor slowing in BD. Reduced DKI metrics in connections traversing motor regions were observed in the current study, consistent with altered connectivity in areas that may contribute to psychomotor slowing in BD. In addition to a role in motor control, the cerebellum is thought to be involved in affect and cognition.^50^ As several pathways emanating from the cerebellum terminate in various frontal and limbic regions, white matter alterations in the cerebellum might contribute to mood dysregulation in BD, which has been linked to disruption in fronto-limbic interconnections.^4^

### Limitations

There are a number of limitations in the present study. First, while the sample size was comparable to other studies of this kind,^42,43^ a larger sample size would have afforded increased power to detect associations between anatomical and clinical measures. Second, similar to most prior studies in this field, our sample was heterogenous with respect to medications and clinical history. Third, as with DTI, there are inherent challenges associated with DKI that limit interpretation of the findings. Similar to DTI, the DKI model is phenomenological and not explicitly based on any single underlying biophysical microstructural paradigm. This means that although DKI is highly sensitive to microstructural differences, we do not know the specific differences in microstructural features (e.g., myelin, axons) that underlie the DKI alterations in BD patients. In line with this, there is a lack of direct association between the streamlines reconstructed through tractography techniques and underlying anatomical and functional connectivity, which confounds neurobiological interpretation of our connectivity findings.

## Conclusion

BD is associated with differences in DKI metrics in several brain regions, which may reflect altered connectivity across cerebellar, subcortical and cortical substructures. The enhanced sensitivity of DKI over DTI to detect these changes suggests that DKI may prove useful in the investigation of subtle white matter microstructural differences in BD. Further studies are required to elucidate the extent of white matter alterations in BD, as well as the cellular and molecular mechanisms underlying these alterations.

## Data Availability

We do not have permission to share individual data from subjects, but can share data as appropriate.

## Acknowledgments

This study was funded by a Hotchkiss Brain Institute/Pfizer Canada Research Grant through the Pfizer Psychiatry Research Awards. Vina Goghari was supported by a Canadian Institutes of Health Research New Investigator Award.

## References

1. Dupont RM, Jernigan TL, Butters N, et al. Subcortical abnormalities detected in bipolar affective disorder using magnetic resonance imaging: Clinical and neuropsychological significance. Arch Gen Psychiatry. 1990;47(1):55–59. doi:10.1001/archpsyc.1990.01810130057008

2. Pezzoli S, Emsell L, Yip SW, et al. Meta-analysis of regional white matter volume in bipolar disorder with replication in an independent sample using coordinates, T-maps, and individual MRI data. Neurosci Biobehav Rev. 2018;84:162–170. doi:10.1016/j.neubiorev.2017.11.005

3. Hu R, Stavish C, Leibenluft E, Linke JO. White matter microstructure in individuals with and at risk for bipolar disorder: Evidence for an endophenotype from a voxel-based meta-analysis. Biol Psychiatry Cogn Neurosci Neuroimaging. Published online June 20, 2020. doi:10.1016/j.bpsc.2020.06.007

4. Mahon K, Burdick KE, Szeszko PR. A role for white matter abnormalities in the pathophysiology of bipolar disorder. Neurosci Biobehav Rev. 2010;34(4):533–554. doi:10.1016/j.neubiorev.2009.10.012

5. Sariçiçek A, Zorlu N, Yalin N, et al. Abnormal white matter integrity as a structural endophenotype for bipolar disorder. Psychol Med. 2016;46(7):1547–1558. doi:10.1017/S0033291716000180

6. Jones DK, Knösche TR, Turner R. White matter integrity, fiber count, and other fallacies: The do’s and don’ts of diffusion MRI. NeuroImage. 2013;73:239–254. doi:10.1016/j.neuroimage.2012.06.081

7. Abramovic L, Boks MPM, Vreeker A, et al. White matter disruptions in patients with bipolar disorder. Eur Neuropsychopharmacol. 2018;28(6):743–751. doi:10.1016/j.euroneuro.2018.01.001

8. Favre P, Pauling M, Stout J, et al. Widespread white matter microstructural abnormalities in bipolar disorder: Evidence from mega-and meta-analyses across 3033 individuals. Neuropsychopharmacology. 2019;44(13):2285–2293. doi:10.1038/s41386-019-0485-6

9. Mahapatra A, Khandelwal SK, Sharan P, Garg A, Mishra NK. Diffusion tensor imaging tractography study in bipolar disorder patients compared to first-degree relatives and healthy controls. Psychiatry Clin Neurosci. 2017;71(10):706–715. doi:10.1111/pcn.12530

10. Haznedar MM, Roversi F, Pallanti S, et al. Fronto-thalamo-striatal gray and white matter volumes and anisotropy of their connections in bipolar spectrum illnesses. Biol Psychiatry. 2005;57(7):733–742. doi:10.1016/j.biopsych.2005.01.002

11. Versace A, Almeida JRC, Hassel S, et al. Elevated left and reduced right orbitomedial prefrontal fractional anisotropy in adults with bipolar disorder revealed by tract-based spatial statistics. Arch Gen Psychiatry. 2008;65(9):1041–1052. doi:10.1001/archpsyc.65.9.1041

12. Houenou J, Wessa M, Douaud G, et al. Increased white matter connectivity in euthymic bipolar patients: diffusion tensor tractography between the subgenual cingulate and the amygdalo-hippocampal complex. Mol Psychiatry. 2007;12(11):1001–1010. doi:10.1038/sj.mp.4002010

13. Phillips ML, Ladouceur CD, Drevets WC. A neural model of voluntary and automatic emotion regulation: implications for understanding the pathophysiology and neurodevelopment of bipolar disorder. Mol Psychiatry. 2008;13(9):833–857. doi:10.1038/mp.2008.65

14. Koshiyama D, Fukunaga M, Okada N, et al. White matter microstructural alterations across four major psychiatric disorders: mega-analysis study in 2937 individuals. Mol Psychiatry. 2020;25(4):883–895. doi:10.1038/s41380-019-0553-7

15. Poletti S, Melloni E, Aggio V, et al. Grey and white matter structure associates with the activation of the tryptophan to kynurenine pathway in bipolar disorder. J Affect Disord. 2019;259:404–412. doi:10.1016/j.jad.2019.08.034

16. Ambrosi E, Chiapponi C, Sani G, et al. White matter microstructural characteristics in Bipolar I and Bipolar II Disorder: A diffusion tensor imaging study. J Affect Disord. 2016;189:176–183. doi:10.1016/j.jad.2015.09.035

17. Lan MJ, Rubin-Falcone H, Sublette ME, et al. Deficits of white matter axial diffusivity in bipolar disorder relative to major depressive disorder: No relationship to cerebral perfusion or body mass index. Bipolar Disord. 2020;22(3):296–302. doi:10.1111/bdi.12845

18. Haller S, Xekardaki A, Delaloye C, et al. Combined analysis of grey matter voxel-based morphometry and white matter tract-based spatial statistics in late-life bipolar disorder. J Psychiatry Neurosci JPN. 2011;36(6):391–401. doi:10.1503/jpn.100140

19. Jeurissen B, Leemans A, Tournier J, Jones DK, Sijbers J. Investigating the prevalence of complex fiber configurations in white matter tissue with diffusion magnetic resonance imaging. Hum Brain Mapp. 2013;34(11):2747–2766. doi:10.1002/hbm.22099

20. Jensen JH, Helpern JA. MRI quantification of non-Gaussian water diffusion by kurtosis analysis1. NMR Biomed. 2010;23(7):698–710. doi:10.1002/nbm.1518

21. Helpern JA, Adisetiyo V, Falangola MF, et al. Preliminary evidence of altered gray and white matter microstructural development in the frontal lobe of adolescents with attention-deficit hyperactivity disorder: A diffusional kurtosis imaging study. J Magn Reson Imaging. 2011;33(1):17–23. doi:10.1002/jmri.22397

22. Tournier J-D, Calamante F, Connelly A. Robust determination of the fibre orientation distribution in diffusion MRI: Non-negativity constrained super-resolved spherical deconvolution. NeuroImage. 2007;35(4):1459–1472. doi:10.1016/j.neuroimage.2007.02.016

23. Michael B. First, Janet B.W. Williams, Rhonda S. Karg, Robert L. Spitzer. Structured Clinical Interview for DSM-5 -Research Version (SCID-5 for DSM-5, Research Version: SCID-5-RV, Version 1.0.0). American Psychiatry Association; 2015.

24. Young RC, Biggs JT, Ziegler VE, Meyer DA. A rating scale for mania: Reliability, validity and sensitivity. Br J Psychiatry. 1978;133(5):429–435. doi:10.1192/bjp.133.5.429

25. Williams JBW. A structured interview guide for the Hamilton Depression Rating Scale. Arch Gen Psychiatry. 1988;45(8):742. doi:10.1001/archpsyc.1988.01800320058007

26. Rosa AR, Sánchez-Moreno J, Martínez-Aran A, et al. Validity and reliability of the Functioning Assessment Short Test (FAST) in bipolar disorder. Clin Pract Epidemiol Ment Health CP EMH. 2007;3:5–5. doi:10.1186/1745-0179-3-5

27. Bruce Rybarczyk. Social and occupational functioning assessment scale (SOFAS). In: Encyclopedia of Clinical Neuropsychology (Kreutzer J, DeLuca J, Caplan B, Eds). Springer; 2011:2313.

28. David Wechsler. Wechsler Test of Adult Reading: WTAR. Psychological Corporation; 2001.

29. Mark Jenkinson, Christian F. Beckmann, Timothy E. J. Behrens, Mark W. Woolrich, Stephen M. Smith. Fsl. Vol 62. Academic Press; 2012.

30. Andersson JLR, Skare S, Ashburner J. How to correct susceptibility distortions in spin-echo echo-planar images: Application to diffusion tensor imaging. NeuroImage. 2003;20(2):870–888. doi:10.1016/S1053-8119(03)00336-7

31. Andersson JLR, Sotiropoulos SN. An integrated approach to correction for off-resonance effects and subject movement in diffusion MR imaging. NeuroImage. 2016;125(Complete):1063–1078. doi:10.1016/j.neuroimage.2015.10.019

32. Leemans A, Jones DK. The B-matrix must be rotated when correcting for subject motion in DTI data. Magn Reson Med. 2009;61(6):1336–1349. doi:10.1002/mrm.21890

33. Smith SM. Fast robust automated brain extraction. Hum Brain Mapp. 2002;17(3):143–155. doi:10.1002/hbm.10062

34. Leemans A, Jeurissen B, Sijbers J, Jones DK. ExploreDTI: A graphical toolbox for processing, analyzing, and visualizing diffusion MR data. :1.

35. Jensen JH, Helpern JA, Ramani A, Lu H, Kaczynski K. Diffusional kurtosis imaging: The quantification of non-gaussian water diffusion by means of magnetic resonance imaging. Magn Reson Med. 2005;53(6):1432–1440. doi:10.1002/mrm.20508

36. Veraart J, Sijbers J, Sunaert S, Leemans A, Jeurissen B. Weighted linear least squares estimation of diffusion MRI parameters: Strengths, limitations, and pitfalls. NeuroImage. 2013;81:335–346. doi:10.1016/j.neuroimage.2013.05.028

37. Klein S, Staring M, Murphy K, Viergever MA, Pluim J. Elastix: A toolbox for intensity-based medical image registration. IEEE Trans Med Imaging. 2010;29(1):196–205. doi:10.1109/TMI.2009.2035616

38. Winkler AM, Ridgway GR, Webster MA, Smith SM, Nichols TE. Permutation inference for the general linear model. NeuroImage. 2014;92(Complete):381–397. doi:10.1016/j.neuroimage.2014.01.060

39. Tax CMW, Jeurissen B, Vos SB, Viergever MA, Leemans A. Recursive calibration of the fiber response function for spherical deconvolution of diffusion MRI data. NeuroImage. 2014;86:67–80. doi:10.1016/j.neuroimage.2013.07.067

40. Zalesky A, Fornito A, Bullmore ET. Network-based statistic: Identifying differences in brain networks. NeuroImage. 2010;53(4):1197–1207. doi:10.1016/j.neuroimage.2010.06.041

41. Nichols TE, Holmes AP. Nonparametric permutation tests for functional neuroimaging: A primer with examples. Hum Brain Mapp. 2002;15(1):1–25. doi:10.1002/hbm.1058

42. Ota M, Noda T, Sato N, et al. The use of diffusional kurtosis imaging and neurite orientation dispersion and density imaging of the brain in bipolar disorder. J Affect Disord. 2019;251:231–234. doi:10.1016/j.jad.2019.03.068

43. Sawamura D, Narita H, Hashimoto N, et al. Microstructural alterations in bipolar and major depressive disorders: A diffusion kurtosis imaging study. J Magn Reson Imaging. 2020;52(4):1187–1196. doi:10.1002/jmri.27174

44. Emsell L, Leemans A, Langan C, et al. Limbic and callosal white matter changes in euthymic bipolar I disorder: An advanced diffusion magnetic resonance imaging tractography study. Biol Psychiatry. 2013;73(2):194–201. doi:10.1016/j.biopsych.2012.09.023

45. Forde NJ, O’Donoghue S, Scanlon C, et al. Structural brain network analysis in families multiply affected with bipolar I disorder. Psychiatry Res Neuroimaging. 2015;234(1):44–51. doi:10.1016/j.pscychresns.2015.08.004

46. Wu EX, Cheung MM. MR diffusion kurtosis imaging for neural tissue characterization1. NMR Biomed. 2010;23(7):836–848. doi:10.1002/nbm.1506

47. Lu LH, Zhou XJ, Fitzgerald J, et al. Microstructural abnormalities of white matter differentiate pediatric and adult-onset bipolar disorder. Bipolar Disord. 2012;14(6):597–606. doi:10.1111/j.1399-5618.2012.01045.x

48. Zhao L, Wang Y, Jia Y, et al. Cerebellar microstructural abnormalities in bipolar depression and unipolar depression: A diffusion kurtosis and perfusion imaging study. J Affect Disord. 2016;195:21–31. doi:10.1016/j.jad.2016.01.042

49. Bracht T, Federspiel A, Schnell S, et al. Cortico-cortical white matter motor pathway microstructure is related to psychomotor retardation in major depressive disorder. PLOS ONE. 2012;7(12):e52238. doi:10.1371/journal.pone.0052238

50. Schmahmann JD, Guell X, Stoodley CJ, Halko MA. The theory and neuroscience of cerebellar cognition. Annu Rev Neurosci. 2019;42(1):337–364. doi:10.1146/annurev-neuro-070918-050258

