## Supplementary Figure 1 for "Diffusion kurtosis imaging of white matter in bipolar disorder"

**Supplementary Material**

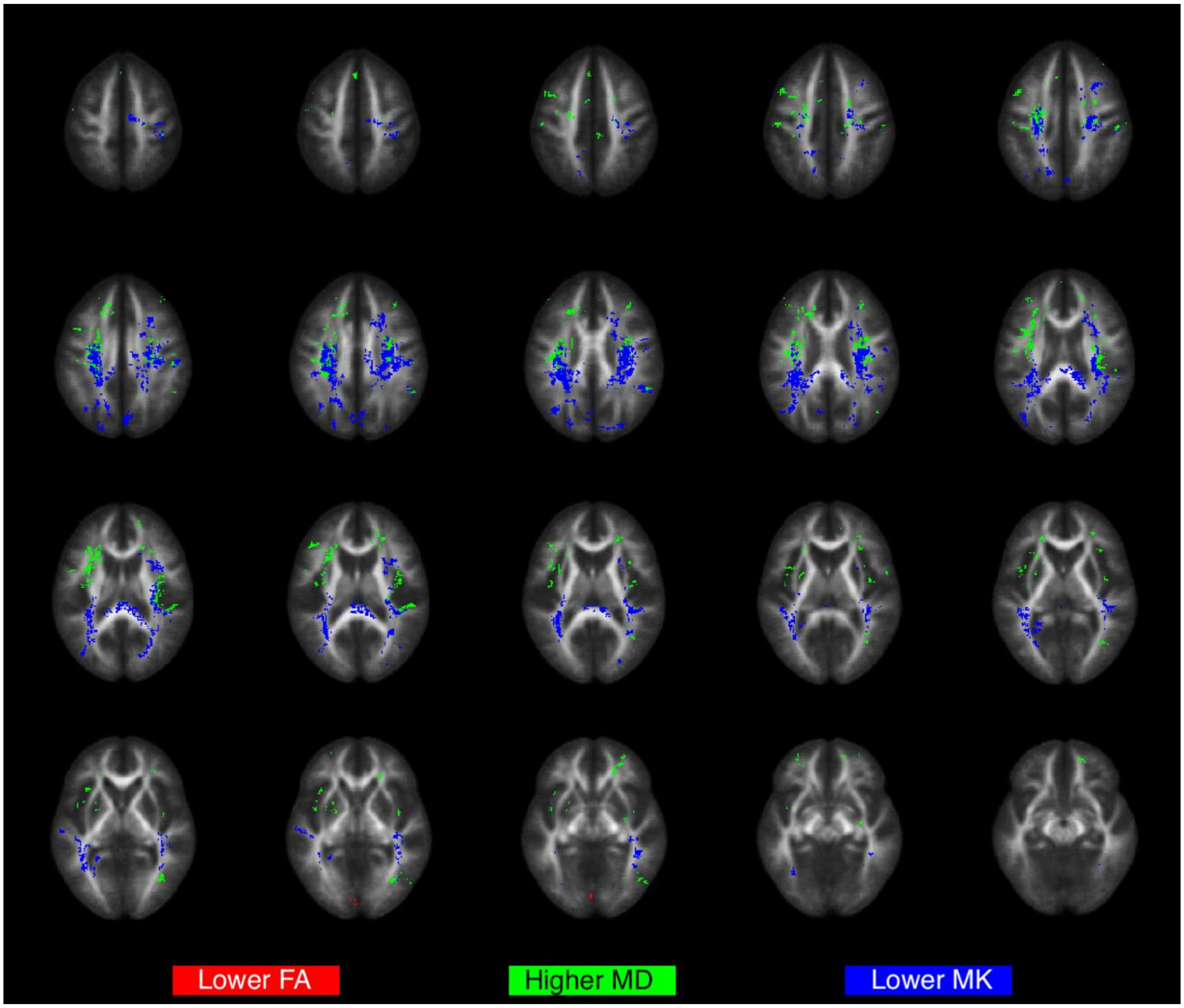


**Supplementary Figure 1: Voxel-wise differences in FA, MD, and MK between bipolar patients and controls.** The image shows voxel-wise differences in FA, MD and MK between the two groups when thresholding the t-test probability at $p < 0.2$ after TFCE correction. Lowering the significance threshold resulted in larger difference clusters for MK, and additional differences in MD, with minor differences in FA.
